# Filovirus Evidence Maps: A community resource to identify and curate the published evidence on immunity and vaccination for BDBV, EBOV, MARV, and SUDV

**DOI:** 10.64898/2026.05.22.26353826

**Authors:** Yuna Chung, Bronwyn A Bailey, Ellen Bowden-Reid, Mariah Csolle, Steffen S Docken, Kim Jachno, David S Khoury, Steve McDonald, Loyal Pattuwage, Heath White, Tsharni Zazryn, Tari Turner, Miles P Davenport

## Abstract

Filoviruses pose a threat to individuals and the global community as pathogens of pandemic potential. The scientific community faces an ongoing challenge of developing effective vaccines with unpredictable outbreaks concentrated in countries with lower healthcare resources. Given these limitations, it is important to ensure that existing filovirus research is used as efficiently as possible. To enable rapid identification and use of this research, we have developed evidence maps of existing filovirus publications to enable further analysis and synthesis. We systematically identified and categorised existing immunological and clinical publications on Bundibugyo (BDBV), Marburg (MARV), Sudan (SUDV) and Ebola (EBOV) viruses. We captured studies that reported on animal or human immune responses to infection, outcome of infection, or human vaccine safety data. Initial searches of PubMed, Embase and Europe PMC were run between November 2024 and January 2025 and the MARV, SUDV and EBOV searches were updated on 1 August 2025. A BDBV search was conducted on 18 May 2026 in response to the WHO declaration of a Public Health Emergency on 17 May 2026. The initial searches retrieved 208, 1646, 534 and 3963 manuscripts for BDBV, MARV, SUDV and EBOV, respectively. After screening using an ‘a priori’ exclusion criteria, 49 BDBV, 198 MARV, 149 SUDV and 850 EBOV publications were included on each evidence map. These maps provide a comprehensive, transparent and reproducible structure to categorise existing studies of filovirus vaccination and immunity. They allow rapid identification of the totality of available evidence and the existing experimental tools to support vaccine development for these priority pathogens.

## Background

On 17 May 2026 the outbreak of Ebola disease (caused by Bundibugyo virus, BDBV) was declared a public health emergency of international concern (PHEIC) (1). This follows recent outbreaks of Marburg (MARV), (2) Ebola (EBOV) (3) and Sudan (SUDV) (4) viruses, which demonstrate the ongoing threat filoviruses pose both to local communities and to global health as pathogens of pandemic potential (5). Despite this, global resources to support research underpinning filovirus vaccine development are limited and conduct of research is further complicated by the sporadic nature of outbreaks.

While there are two vaccines prequalified by the World Health Organization (WHO) for Ebola Zaire virus disease, one of these (Zabdeno/Mvabea) recently had the European Medicines Agency marketing authorisation in the EU withdrawn at the request of the manufacturer (6-8). Further, there are currently no approved vaccines for protection against BDBV, SUDV or MARV disease.

Given the recent health emergency caused by BDBV, and the relatively small investment in therapeutic and vaccine development for this virus to-date, it is important to ensure that the existing research for BDBV and the related filoviruses (MARV, EBOV and SUDV) is accessible and can be used as efficiently as possible to support and guide future research for vaccine development. Here, to enable filovirus stakeholders to have a systematic and comprehensive overview of existing research we have generated “immunological evidence maps” to systematically identify and classify the available research relevant to filovirus vaccine development.

Evidence mapping approaches use a comprehensive search to identify all relevant research studies and classify them according to the types of evidence contained; creating a single source of pre-filtered research in an area of interest (9). An evidence map allows researchers, product developers, and policy makers to rapidly access relevant research literature, without conducting their own exhaustive searches. Researchers and funders can use the maps to assess the totality of evidence to identify areas where there is strong evidence to guide decision-making, as well as to identify research areas where there is little or no data (evidence gaps), allowing prioritisation of future research and targeted funding (10).

## Methods

Evidence mapping is an evidence curation technique complementary to systematic review methods. Both methodologies use a systematic strategy to search research databases to identify potentially relevant publications, after which the titles and abstracts are screened using *a priori* inclusion and exclusion criteria to eliminate irrelevant publications. The full text of each article is then reviewed to confirm the article contains relevant information. In evidence maps, relevant articles are ‘tagged’ to indicate the type of information they contain and then displayed visually on a map, classified according to the type(s) of content (11). The resulting evidence maps are presented visually and can be filtered to explore the evidence on a chosen topic.

The application of evidence mapping in immunology has been adapted from its use in clinical research. Initially, the goal of this study was to create three evidence maps that identify, classify and visually display the research literature on EBOV, SUDV and MARV immunity and vaccination. As this manuscript was in preparation, the WHO declared the Bundibugyo (BDBV) virus outbreak in DRC as a PHEIC on 17 May 2026 and the mapping was extended to BDBV (1). The studies in the evidence maps are classified by pathogen, infection model (human or animal), and the types of immunological assays or infection outcome reported.

### Planning and registration

Our study aimed to identify primary studies and systematic reviews that evaluated an animal or human immune response to each of the relevant filovirus pathogens (natural or challenged), or to vaccination. For a study to be included, the outcome reported needed to relate to either a virus-specific immune measure (e.g. antibody or cellular responses measured in immune assays after infection or vaccination), an infection outcome (e.g. survival or viral levels after infection), or human vaccine safety data. There were no restrictions by publication date, but publications were limited to English language only. Preprints were eligible (unless superseded by the final publication), but conference abstracts, dissertations, non-systematic reviews, expert opinion, news, book chapters, and protocol papers were excluded from this review. Systematic reviews were included where they met predefined criteria. The full study eligibility criteria for primary studies and systematic reviews are detailed in the supplement.

The map development methodology was pre-registered on Open Science Framework (OSF) at https://osf.io/89u5f

### Search Methods

A systematic search was undertaken to identify all relevant literature. An information specialist (SM) worked collaboratively with methodologists and immunologists to develop a search strategy for PubMed that combined medical subject headings (MeSH) and free-text terms for each virus with MeSH and free-text terms for immunobridging strategy, *in-vitro*, human challenge, animal models and natural history (the full search strategy for each virus is provided in the supplement). The PubMed search was adapted for Ovid Embase. A search of Europe PMC was also conducted, restricted to preprints only. Initial searches were run on 27 November 2024 (MARV), 16 December 2024 (SUDV) and 20 January 2025 (EBOV) and were used to develop the first versions of the evidence maps. The evidence maps for MARV, SUDV, and EBOV were subsequently updated to include publications until the end of July 2025 – updated searches were run on 1 August 2025 for all three filoviruses. Finally, on 18 May 2026 a new search was conducted to extend the mapping process to include BDBV. After completing the initial filovirus evidence maps, it was decided it was not necessary to include a search of Europe PMC (which was limited to preprints) for BDBV because Embase now extensively covers all the major preprint servers.

Records retrieved from the database searches for each virus were imported to EndNote and duplicates automatically removed. Deduplicated records were imported to Covidence (12) for screening with any additional duplicates removed automatically. Manual identification of duplicates also occurred where the preprint had been superseded by the final publication.

### Study selection and screening

Five authors participated in study selection and screening. The titles and abstracts of the publications retrieved by the search were screened for eligibility independently by two authors (YC, KJ, LP, MC, MPD), using specified inclusion and exclusion criteria (see supplement). Disagreements were resolved through discussion or by consultation with a senior investigator (MPD). Records deemed eligible at the title/abstract screen were then reviewed for eligibility at full text by two of the five reviewers (YC, KJ, LP, MC, MPD). Both title/abstract and full text screening were carried out in Covidence.

### Evidence map framework

The evidence map framework is a two-dimensional matrix with studies classified by ‘population’ and ‘exposure’ in the columns and ‘outcome’ for the rows. The ‘population’ category classifies studies by species (e.g. human, non-human primate (NHP), mouse, etc.), with subclassifications for the type of exposure (i.e. infection challenge, natural history, vaccination). In the rows, immunogenicity outcomes are divided into subcategories by the type of assay used to measure the response (e.g. antibody binding assay, T cell ELISPOT assay). Studies of filovirus infection were also classified by the type of infection outcomes reported (e.g. survival, viral level) or vaccine safety outcomes. The framework was modified during the project in response to internal piloting and stakeholder feedback. See Figure 1 for an outline of final categories.

**Figure 1.**
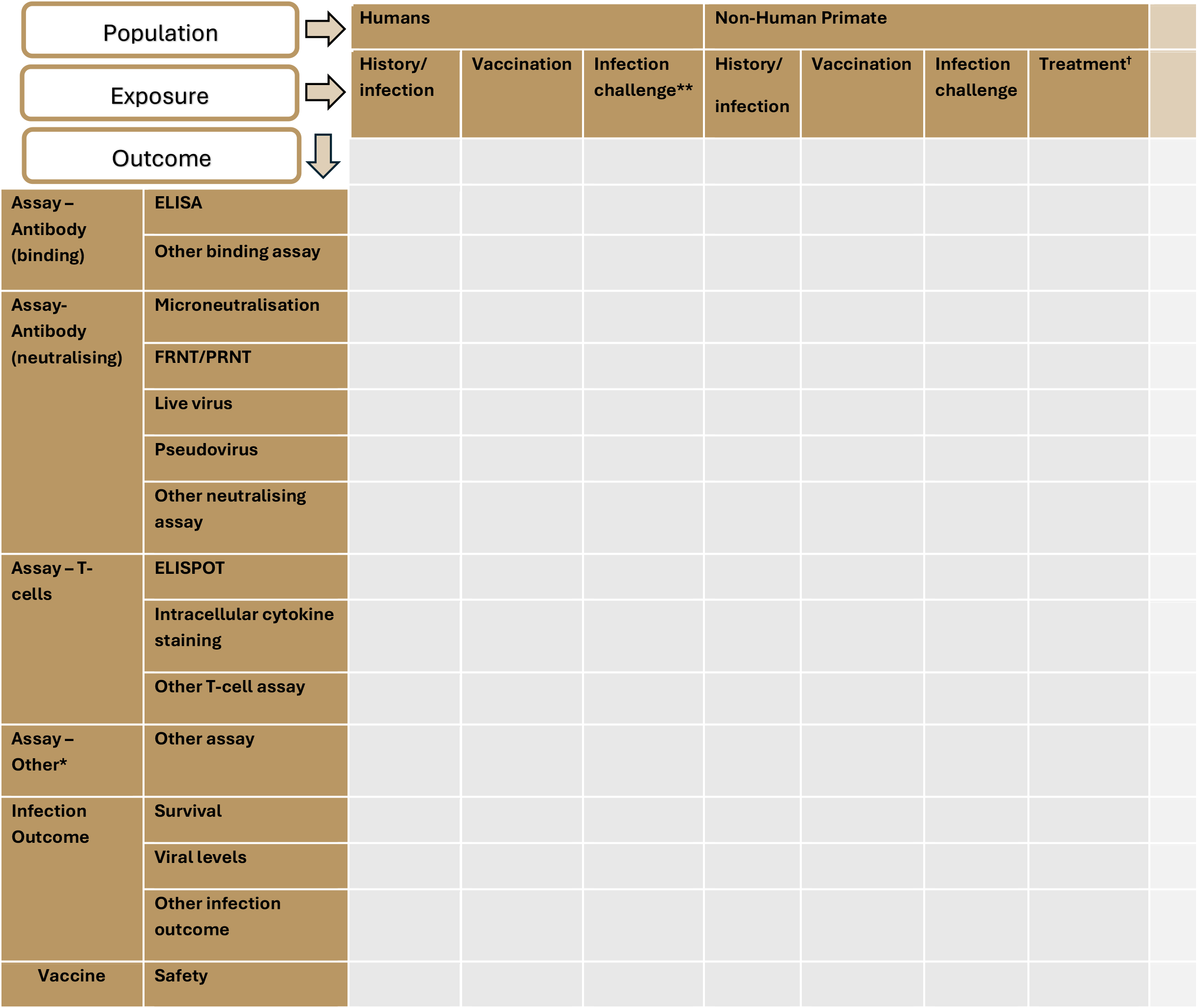
Outline of final evidence map framework. *An additional row is included for ‘other assay’ if an assay does not fit in other listed options. **No infection-challenge studies have been undertaken in humans for filoviruses †Treatment only included studies of antibody treatment (ie: excluded studies of small molecule antivirals)

Each included publication was categorised (‘tagged’) in Covidence according to the populations, exposures, and outcomes by one reviewer and checked by a second reviewer (YC, KJ, LP, MC, MPD). A decision tree was developed to ensure consistent labelling between reviewers (see supplement). Tagging discrepancies between reviewers were resolved by discussion and consultation with the senior investigator (MPD). Many publications had multiple tags (multiple species, exposures, or outcomes reported). As a result, these publications were mapped in multiple cells in the final evidence map. As is standard practice for an evidence map, we did not assess the methodological quality or risk of bias of the included studies, nor did we extract data from the studies.

Following completion of the tagging of studies in Covidence, details from each included study were exported as a csv file. These files were converted to .json format files for display in the evidence mapping software EPPI-Mapper. (13)

### Ethics

This work was approved under the UNSW Sydney Human Research Ethics Committee (approval HC200242).

## Results

Completed maps are available as an interactive webpage on the ALEC website (https://livingevidence.org.au/research-initiatives/about-feeva/evidence-maps/). See Figure 2 for a static visual representation of the BDBV evidence map.

**Figure 2.**
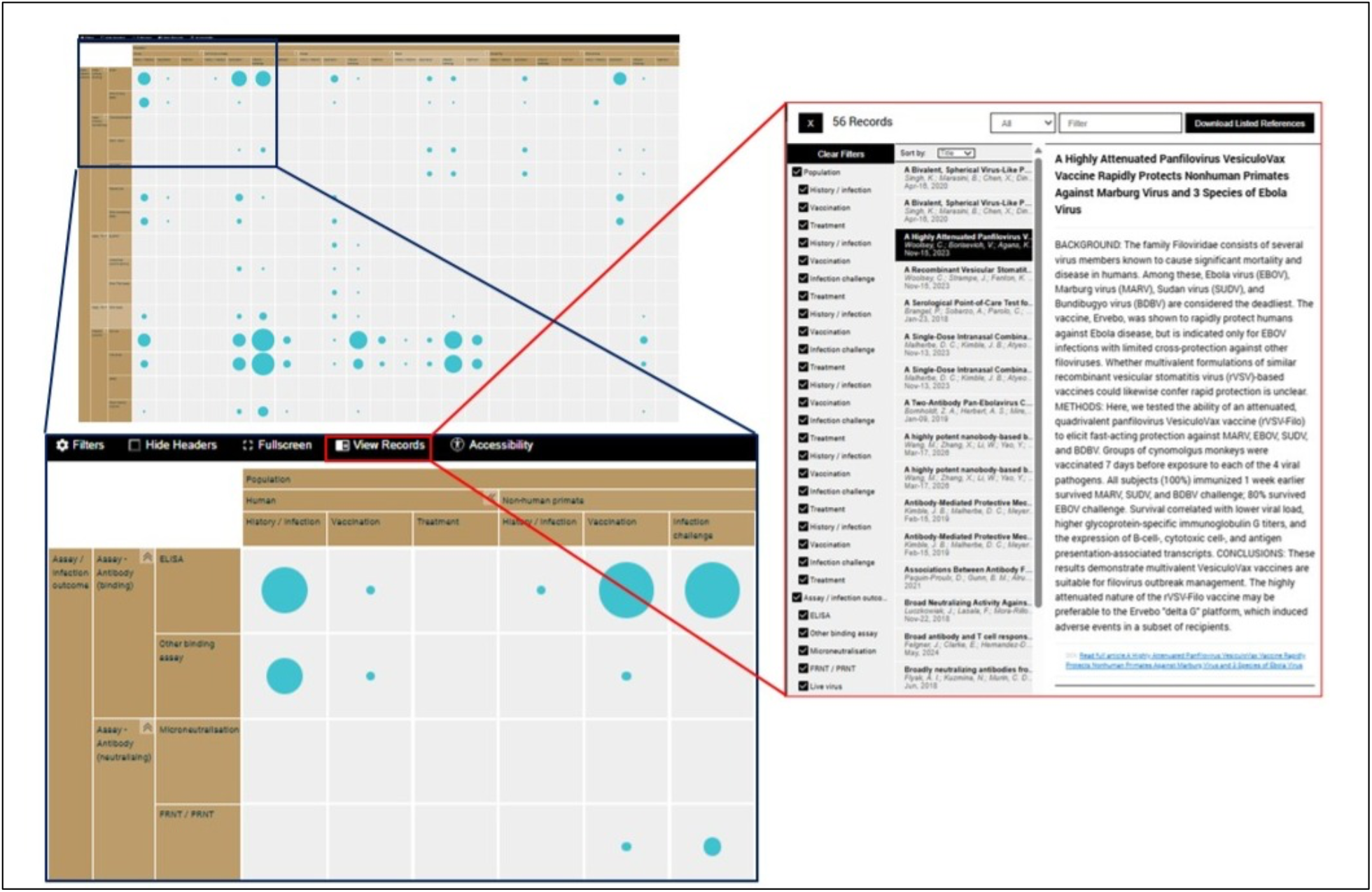
Screenshots of the Bundibugyo Virus evidence map. Studies are categorised by population (human or animal species), exposure, and the immune assays and/or infection outcomes reported.

### Identification and screening of studies

The searches of PubMed, Embase and Europe PMC retrieved 208, 1646, 534 and 3963 records for BDBV, MARV, SUDV and EBOV, respectively. Figure 3 provides a brief outline of the screening process at title/abstract and full-text review. Standard PRISMA flow diagrams for each pathogen are provided in the supplement. Due to the larger volume of studies for EBOV, the systematic reviews search was conducted separately. Of the 127 records retrieved by the search,13 systematic reviews were assessed at full text, with seven of these being included in the map. Overall, after title/abstract screening, almost 30% of publications proceeded to full text review (ranging from 22% for MARV to 44% for SUDV). Approximately 75% of publications (ranging from 60% for MARV to 80% for EBOV) were found to be eligible for tagging and inclusion in the maps following full-text review.

**Figure 3.**
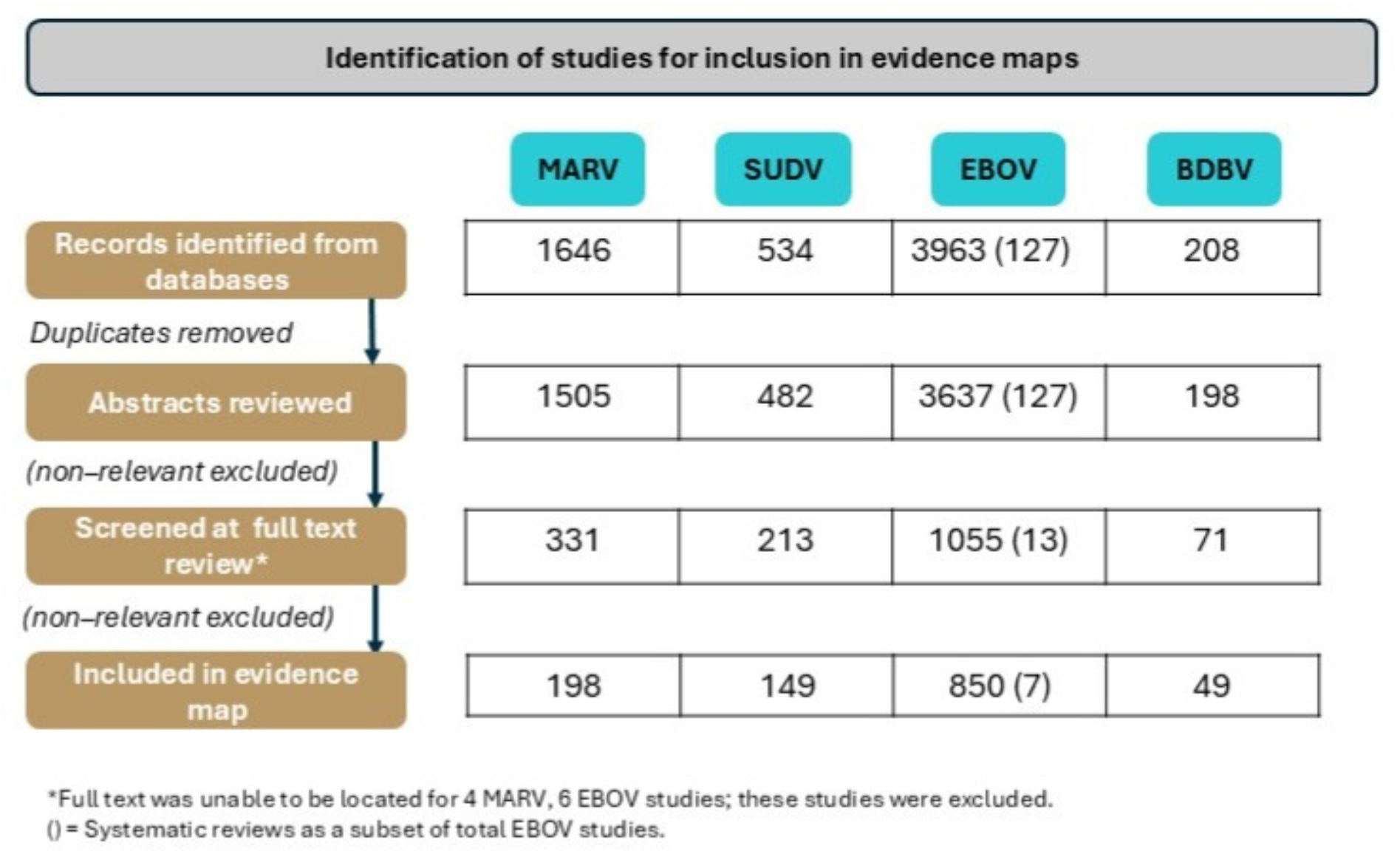
Summary of screening process and study numbers for each filovirus evidence map (full PRISMA flow diagrams available in supplementary materials)

### Mapping the evidence

For all filoviruses, the number of publications in the evidence maps has increased rapidly since the start of this century. The exception is BDBV, as it was initially identified in late 2007 and the first publication included in the map is 2008 (14). The sharp increase in the number of publications in the 2015-2019 period may have been in response to the large West African EBOV outbreak beginning in 2014 (Figure 4) (14). This increase coincided with a large increase in funding for filovirus research and development in 2015 (15). Human data was reported in 20% of MARV, 30% of SUDV, 31% of BDBV and 34% of EBOV publications. Non-human primate data was reported in approximately 37% of MARV, 30% of SUDV, 28% of EBOV, and 25% of BDBV publications. For BDBV, 10 publications included mouse models and 8 included ferret models. Multiple publications reported studies in several species; these publications appear in multiple cells on the map (and therefore percentages add to greater than 100%). Several publications provided results for multiple filoviruses and therefore appear on more than one map.

**Figure 4.**
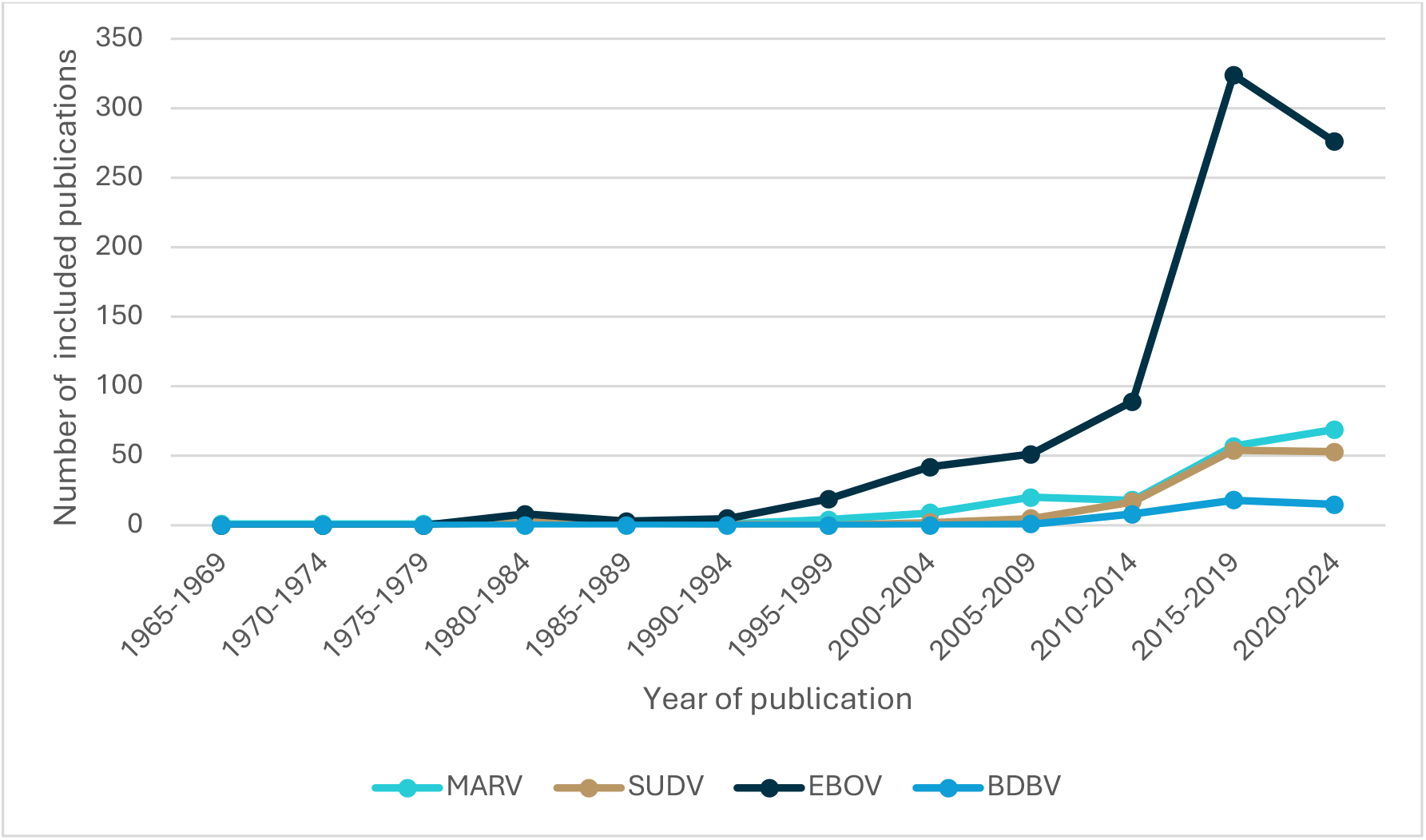
Number of included publications by date of publication (to end 2024)

The most frequently used assay for all filovirus publications was an antibody binding assay (used in 34 BDBV, 132 MARV, 112 SUDV and 552 EBOV publications) followed by *in vitro* neutralisation assays (used in 17 BDBV, 57 MARV, 42 SUDV publications and 259 EBOV publications) (Table 1). T-cell assays were used in 4 BDBV, 47 MARV, 38 SUDV, and 218 EBOV publications. All viruses included publications reporting the use of ‘other’ assay types (e.g., transcriptomic analysis of immune responses, or cytokine and chemokine levels). The number of publications that have included a specific assay does not give any insight into the suitability of the assay type as there has been no assessment of the quality of the publications or the extent to which the use of the assay has been described.

**Table 1.** Number of publications by assay for each population group*.

|  | Human | NHP | Mouse | Ferret | Guinea Pig | Other animal |
| --- | --- | --- | --- | --- | --- | --- |
| **Antibody binding assays (e.g. ELISA)** |  |  |  |  |  |  |
| MARV | 32 | 46 | 22 | 1 | 22 | 21 |
| SUDV | 36 | 31 | 23 | 2 | 11 | 10 |
| EBOV | 213 | 128 | 151 | 3 | 48 | 45 |
| BDBV | 10 | 8 | 4 | 2 | 2 | 8 |
| **Antibody neutralising assays (e.g. PRNT)** |  |  |  |  |  |  |
| MARV | 9 | 22 | 11 | 1 | 11 | 6 |
| SUDV | 9 | 10 | 11 | 2 | 6 | 5 |
| EBOV | 86 | 58 | 79 | 2 | 25 | 21 |
| BDBV | 3 | 4 | 2 | 2 | 2 | 4 |
| **T-cell assays (e.g. ELISPOT)** |  |  |  |  |  |  |
| MARV | 6 | 21 | 16 | 0 | 3 | 1 |
| SUDV | 11 | 10 | 16 | 0 | 0 | 1 |
| EBOV | 57 | 56 | 99 | 1 | 6 | 2 |
| BDBV | 0 | 2 | 2 | 0 | 0 | 0 |
| **Other assays** |  |  |  |  |  |  |
| MARV | 7 | 39 | 13 | 0 | 10 | 14 |
| SUDV | 10 | 8 | 6 | 0 | 4 | 0 |
| EBOV | 51 | 60 | 62 | 4 | 5 | 4 |
| BDBV | 2 | 3 | 2 | 1 | 0 | 1 |

\* NB. The total number of publications in the cells adds to greater than the number of publications for each map as some publications are included in multiple cells. A higher number of studies in a cell within a map does not necessarily indicate strength or quality of evidence.

An infection outcome was reported after either natural exposure or following experimental challenge (animal models only). Survival was the most frequently measured infection outcome (considering all species, including humans) across all pathogens, reported in 31, 110, 81 and 447 for BDBV, MARV, SUDV and EBOV publications, respectively. Non-human primate models had the highest number of studies with survival as an outcome for BDBV, MARV and SUDV, whereas mouse models were the most common for EBOV. Viral levels were reported as an infection outcome for 25 BDBV, 110 MARV, 55 SUDV and 359 EBOV publications. Viral levels were reported most frequently in NHP models for all four filoviruses. Comparatively, fewer publications measured vaccine safety or other outcomes for each filovirus. There was no vaccine safety data available for BDBV (Table 2). A higher number of studies reporting an outcome does not indicate that the evidence is more reliable as no assessment of the quality of the evidence was undertaken when developing the evidence maps. An assessment of evidence quality is the next step for users of the maps when exploring a specific research question.

**Table 2.**
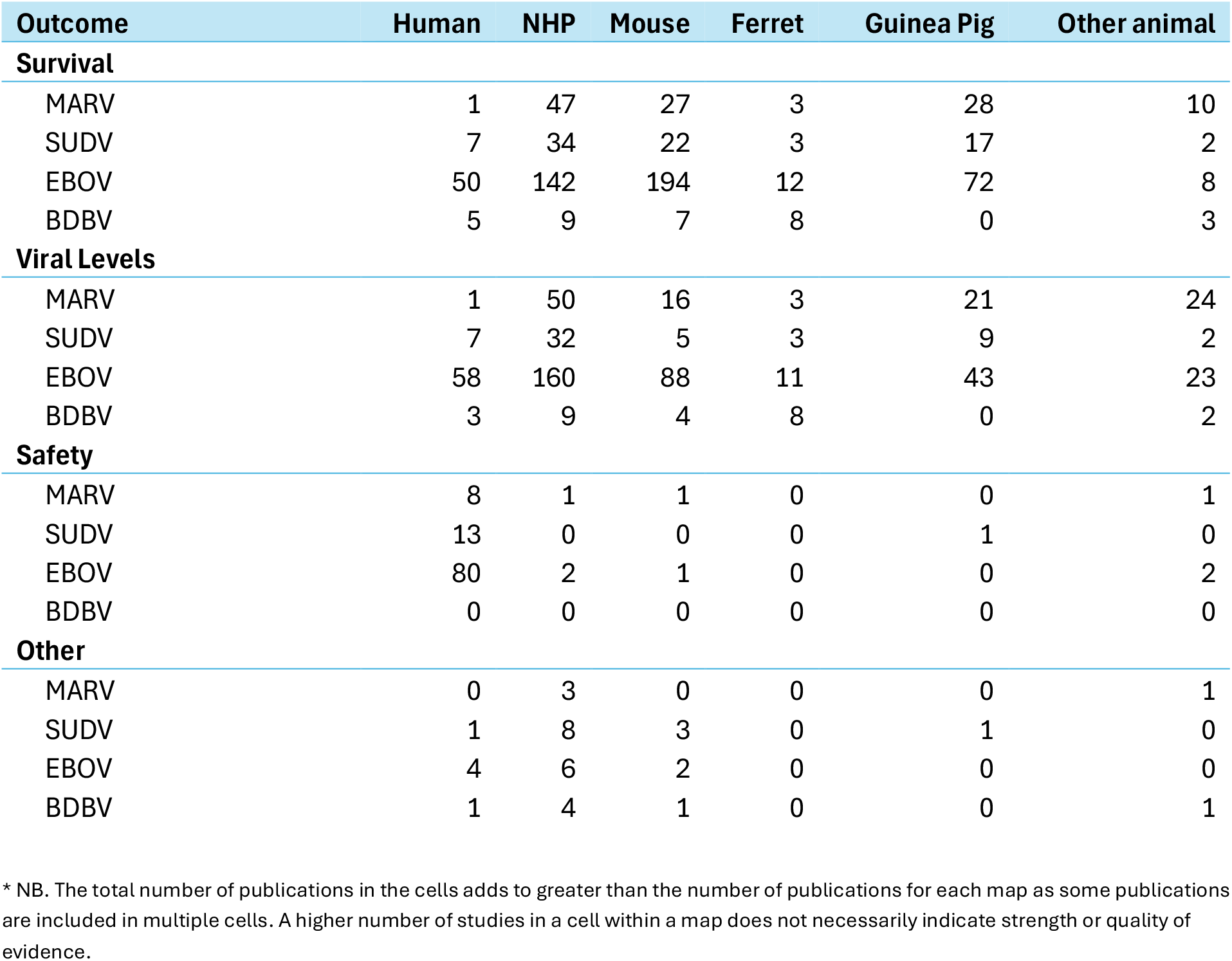
Number of publications by infection outcome for each population group*.

The rows in table 1 and 2 may not add to the publication’s results in the text above, as some publications may have included studies with two or more animal models. Many publications reported the use of multiple assays or outcomes, so the total number of publications for each pathogen in table 1 and 2 is greater than the number of publications in the maps.

## Discussion

The current public health emergency in the DRC and Uganda highlights the need for accelerating the development, assessment and approval of an effective vaccine and other medical countermeasures for BDBV. Further, major outbreaks of filoviruses are an ongoing and increasing global health concern (14). Therefore, the evidence maps here are not only aimed to provide a resource for each individual pathogen, but the four filovirus maps also allow rapid analysis of what has been achieved in similar studies across-pathogens. For example, the original licensure of the Zabdeno / Mvabea vaccine against EBOV by the European Medicines Agency (EMA) (6, 7) was supported by data on antibody binding and protection in NHP, and antibody binding after vaccination in human clinical trials. To assess the potential for a similar pathway in Bundibugyo, a researcher may choose to review all NHP vaccination and challenge studies with BDBV, as well as the assays used to measure antibody responses in these studies. These NHP infection models and antibody binding assays can also be compared with those used in Ebola by reference to the Ebola evidence map. In the case of Bundibugyo, the maps can be used to identify gaps in the current evidence landscape where new primary research may be required to provide evidence for immunity or vaccine effectiveness, avoiding duplication, and focusing future research efforts.

Systematic reviews of the available evidence to address particular questions are of enormous value, and in cases with limited investment can leverage the existing research to identify new insights (16). A small number of systematic reviews have been performed on filovirus immunity, for example summarising the results of clinical and NHP studies of immunity (17, 18). However, this approach requires each systematic reviewer to conduct exhaustive searches and study selection for each specific research question. Evidence maps can greatly accelerate this process by aggregating and classifying the available literature across a broad topic area. They can operate as the first step in a systematic review for multiple filovirus research questions and remove the need for individual investigators to search the literature separately to identify relevant studies. Researchers can focus on the studies in the map cells relevant to their question and focus their efforts on appraising and synthesising the studies within these cells. In addition to aggregating the available evidence, these maps will ideally be used by funders and vaccine developers to identify gaps where key studies should be prioritized. For example, evidence on human convalescent immune responses and vaccine immunogenicity for BDBV are currently lacking, and the evidence on any association between antibody binding and vaccine protection in NHP is sparse.

Regularly updating evidence syntheses (i.e. making them ‘living’) is important for research areas like filoviruses, where research evidence is emerging, current evidence is uncertain or where new research might change policy or practice. The value of updating evidence synthesis and guidelines is increasingly accepted in clinical research (19). To ensure the maps continue to accurately reflect the research landscape, the researchers recommend continuing to update the evidence maps with new studies every six months, and perhaps more regularly during PHEIC such as the present one. To date, one update to include new literature of the evidence maps has been completed (outlined in the methods) for EBOV, MARV and SUDV.

An important limitation of evidence maps is that a higher number of publications in a given cell within a map does not necessarily indicate a high quantity or quality of evidence. It can be that (for example) a large number of very small studies, or studies lacking necessary controls, may be found in a given cell of the map and which do not provide the required evidence or quality of evidence to address a given question. How immunological evidence can be evaluated for quality is also area that requires ongoing investment (20).

Evidence mapping is a comparatively new research methodology, and has most often been applied to clinical literature (21). To our knowledge, these are the first evidence maps categorising studies reporting measures of immune response to infection or vaccination. The application of evidence mapping approaches to preclinical research presents several challenges. Firstly, clinical studies often include structured abstracts and standardised reporting tools that allow easy identification of the relevant study details. By contrast, immunological studies in animals often include narrative abstracts and limited reporting of experimental procedures. Standardised reporting tools such as the ARRIVE guidelines 2.0 (22) attempt to address this issue by identifying the minimum information necessary to report in publications describing *in vivo* experiments, however these approaches are not universally applied. More structured, detailed, and standardised reporting would substantially improve the efficiency of mapping of preclinical studies.

Another major challenge is that preclinical publications often present multiple research studies in different animal models and with more than one outcome and/or measurement tool. This can make it difficult to precisely map an individual publication. As an example, a manuscript may include studies of ELISPOT numbers after vaccination in NHPs, as well as a separate study where NHPs were infected and only the infection outcome (survival) was reported. Due to the limitations of existing evidence mapping software, for this publication both the columns (‘vaccination’ and ‘infection challenge’) would appear to intersect both the rows (e.g. ELISPOT, Outcome survival) – even though ELISPOT was not reported in the infection challenge study and survival was not reported in the vaccination study. Thus, it is important to recognise that the goal is to include all the publications relevant to a specific cell on the map, so inclusivity was prioritised. This ensures that all research relevant to answering a question will be available in a cell on the map. However, occasionally, some of the publications listed may include non-relevant studies (for the reasons above). We acknowledge that the trade-off for this is that a cell may require further manual filtering by the user to find the research that addresses their question.

Artificial Intelligence (AI) was not used to create the evidence maps beyond what is already available in the Covidence® Systematic review Software, which includes some automation features to increase the speed of the screening phase of evidence review (23). AI and automation may ultimately play a role in accelerating evidence syntheses. However, it is essential to ensure the AI system or tool used is methodologically sound and will not undermine the reliability of the synthesis (24). Importantly, for this work, we could find no examples of how AI has been validated or piloted to ensure that it is appropriate for use in this relatively innovative area of immunological evidence synthesis (24). We recommend a direct comparison of the human curated versus AI developed map to validate the use of AI tools and to assess their readiness to be employed for future pathogen evidence mapping or updates. Moreover, it is worth noting that with the infrastructure in place, even without AI tools, the Bundibugyo evidence map could be generated rapidly because of the existing pre-registered methods, limited literature, and experienced reviewers. That is, the PHEIC was declared on 17 May 2026, and the generation of the map of available evidence on immunity and vaccination was completed by 20 May.

Filoviruses pose an ongoing threat both to local communities and globally as pathogens of pandemic potential. These evidence maps provide the filovirus community with a curated overview of preclinical immunological research for BDBV, MARV, SUDV and EBOV, to help inform future research priorities and define key questions for more focused evidence synthesis. The evidence maps will also allow researchers to confidently assess the totality of available evidence on their research topic, as well as facilitate future evidence synthesis and meta-analysis. Ultimately, these maps provide a resource to support the research, development and deployment of effective vaccines and treatments for filoviruses.

## Data Availability

All data generated in this study are available at https://livingevidence.org.au/research-initiatives/about-feeva/evidence-maps/.

https://osf.io/89u5f

## Funding

This project was supported by funding from The Coalition for Epidemic Preparedness Innovations (CEPI) and PATH. Professor Miles Davenport was supported by NHMRC Investigator Grant GNT2034108. Dr David Khoury was supported NHMRC Investigator Grant GNT2033318.

## Conflicts of Interest

The authors declare no conflicts of interest.

## Acknowledgments

Thank you to the filovirus experts who provided stakeholder feedback on the evidence map as part of the living update.

## Notes

### Competing Interest Statement

The authors have declared no competing interest.

### Author Declarations

This work was approved under the UNSW Sydney Human Research Ethics Committee (approval HC200242).

