## Supplemental Files for "Filovirus Evidence Maps: A community resource to identify and curate the published evidence on immunity and vaccination for BDBV, EBOV, MARV, and SUDV"

*Chung et al.*

### Supplementary materials

#### Filovirus evidence map: inclusion/exclusion criteria.

| **Inclusion Criteria** | **Exclusion Criteria** |
|---|---|
| **Population**<br>- animal (any)<br>- human<br>- serum from animal or human<br><br>**Intervention/Exposure**<br>- vaccines<br>- natural infection<br>- infection challenge/lethal challenge<br>- monoclonal antibody (mAbs) treatment<br><br>**Comparator** (if present)<br>- unvaccinated<br>- untreated<br>- uninfected<br><br>**Outcome**<br>- Binding antibodies<br> - binding ELISA<br> - binding-ELISA OD<br> - binding-ELISA endpoint<br> - binding mean fluorescence intensity (MFI))<br>- Neutralising antibodies<br> - Neutralisation – PRNT/FRNT<br> - Microneutralisation<br>- T cell<br> - T cell ELISPOT<br> - T cell ICS<br> - T cell IGRS<br> - T cell other<br> - CD4<br> - CD8 | - Other filovirus studies that did not report on the specific filovirus of the relevant map<br>- Optimising diagnostic assays for detection of viral load<br>- In vitro assays testing drugs/small molecules<br>- Characterisation of monoclonal antibodies<br>- Generation of monoclonal antibodies in animal model<br><br>Study types<br>- Prevalence/epidemiological studies<br>- Modelling studies<br>- Case-studies<br>- Cost-effectiveness studies<br>- Narrative reviews, editorials, perspectives, commentaries<br>- Book chapters<br>- News articles<br>- Conference abstracts or dissertations<br>- Protocols |

| |
|---|
| - B cell<br>- Infection<br> - viremia<br> - viral load<br>- Survival/death<br>- Vaccine safety (humans)<br><br>Study characteristics:<br>- Marburg/Sudan/Ebola/Bundibugyo virus study types the one that matches the map)<br>- Assays for detection of antibodies in animal or human sera<br>- Systematic reviews |

#### Database search strategies

### Marburg virus (MARV)

Search date: 27 November 2024

**PubMed**

| # | Marburg virus | Search statement | Results |
|---|---|---|---|
| 1 | MeSH Terms and free-text terms for Marburg virus | Marburg Virus Disease[mh] OR Marburgvirus[mh] OR marburg*[ti] OR marburgvirus*[tiab] OR "marburg virus"[tiab:~5] OR "marburg viruses"[tiab:~5] OR "marburg filovirus"[tiab:~5] OR "marburg filoviruses"[tiab:~5] OR "marburg fever"[tiab:~5] OR "marburg fevers"[tiab:~5] OR MARV[tiab] OR marburg*[ot] | 2200 |
| 2 | MeSH Terms and free-text terms for vaccines | Vaccines[mh] OR Vaccine Development[mh] OR Vaccination[mh] OR vaccin*[tiab] | 525,328 |
| 3 | MeSH Terms and free-text terms for immunobridging strategy, in-vitro, human challenge, animal models, natural history | Disease Models, Animal[mh] OR Enzyme-Linked Immunosorbent Assay[mh] OR Antibodies, Neutralizing[mh] OR Antibodies, Monoclonal[mh] OR Neutralization Tests[mh] OR Biological Assay[mh] OR Enzyme-Linked Immunospot Assay[mh] OR Immunity[mh] OR ELISA*[tiab] OR immuno*[tiab] OR immune*[tiab] OR PRNT[tiab] OR neutrali*[tiab] OR monoclonal[tiab] OR mAbs[tiab] OR antiviral*[tiab] OR assay*[tiab] OR luminex[tiab] OR ELISPOT*[tiab] OR T-cell*[tiab] OR "animal model"[tiab:~5] OR "animal models"[tiab:~5] OR non-human*[tiab] OR nonhuman*[tiab] OR primate*[tiab] OR chimpanzee*[tiab] OR macaque*[tiab] OR mice[tiab] OR mouse[tiab] OR hamster*[tiab] OR ferret*[tiab] OR murine[tiab] OR "guinea pig*"[tiab] OR challenge[tiab] OR "natural history"[tiab] OR study[ti] OR trial[ti] | 8,439,682 |
| 4 | Combine sets | #1 AND (#2 OR #3) | 1140 |

**Ovid Embase**

Embase Classic + Embase <1947 to 2024 November 25>

| # | Search Statement | Results |
|---|---|---|
| 1 | Marburgvirus/ or Marburg Hemorrhagic Fever/ or (marburgvirus* or (marburg adj5 virus*) or (marburg adj5 filovirus*) or (marburg adj5 fever*) or MARV).ti,ab. or marburg*.ti. or marburg*.kf,kw. | 2898 |
| 2 | exp Vaccine/ or Vaccine Development/ or Vaccination/ or vaccin*.ti,ab,kf. | 729,228 |
| 3 | Disease Model/ or exp Enzyme Linked Immunosorbent Assay/ or exp Neutralizing Antibody/ or exp Neutralization Test/ or exp bioassay/ or Enzyme Linked Immunospot Assay/ or (ELISA* or immuno* or immune* or PRNT or neutrali* or monoclonal or mAbs or antiviral* or assay* or luminex or ELISPOT* or T-cell* or (animal adj5 model*) or non-human* or nonhuman* or primate* or chimpanzee* or macaque* or mice or mouse or hamster* or ferret* or murine or guinea pig* or human challenge or natural history).ti,ab. or (study or trial).ti. | 10,368,016 |
| 4 | 1 and (2 or 3) | 1516 |
| 5 | exp Review/ or Editorial.pt. | 4,233,595 |
| 6 | 4 not 5 | 1222 |
| 7 | limit 6 to (english language and "remove medline records") | 287 |

##### Europe PMC

Restricted to preprints

((TITLE:"marburg*") OR (TITLE:"MARV") OR (ABSTRACT:"marburg*") OR (ABSTRACT:"MARV")) AND (SRC:PPR) = 91

#### Sudan virus (SUDV)

Search date: 16 December 2024

**PubMed**

| # | Concept | Search statement | Results |
|---|---|---|---|
| 1 | MeSH Terms and free-text terms for Sudan virus (excl Sudan fever/s) | sudanvirus*[tiab] OR "sudan virus"[tiab:~5] OR "sudan viruses"[tiab:~5] OR "sudan ebola"[tiab:~5] OR "sudan ebolavirus"[tiab:~5] OR "sudan ebolaviruses"[tiab:~5] OR "sudan filovirus"[tiab:~5] OR "sudan filoviruses"[tiab:~5] OR SUDV[tiab] | 528 |
| 2 | MeSH Terms and free-text terms for vaccines | Vaccines[mh] OR Vaccine Development[mh] OR Vaccination[mh] OR vaccin*[tiab] | 526,639 |
| 3 | MeSH Terms and free-text terms for immunobridging strategy, in-vitro, human challenge, animal models, natural history | Disease Models, Animal[mh] OR Enzyme-Linked Immunosorbent Assay[mh] OR Antibodies, Neutralizing[mh] OR Antibodies, Monoclonal[mh] OR Neutralization Tests[mh] OR Biological Assay[mh] OR Enzyme-Linked Immunospot Assay[mh] OR Immunity[mh] OR ELISA*[tiab] OR immuno*[tiab] OR immune*[tiab] OR PRNT[tiab] OR neutrali*[tiab] OR monoclonal[tiab] OR mAbs[tiab] OR antiviral*[tiab] OR assay*[tiab] OR luminex[tiab] OR ELISPOT*[tiab] OR T-cell*[tiab] OR "animal model"[tiab:~5] OR "animal models"[tiab:~5] OR non-human*[tiab] OR nonhuman*[tiab] OR primate*[tiab] OR chimpanzee*[tiab] OR macaque*[tiab] OR mice[tiab] OR mouse[tiab] OR hamster*[tiab] OR ferret*[tiab] OR murine[tiab] OR "guinea pig*"[tiab] OR challenge[tiab] OR "natural history"[tiab] OR study[ti] OR trial[ti] | 8,463,340 |
| 4 | Combine sets | #1 AND (#2 OR #3) | 377 |

#### Ovid Embase

Embase Classic + Embase <1947 to 2024 December 13>

| # | Search Statement | Results |
|---|---|---|
| 1 | (sudanvirus* or (sudan adj5 virus*) or (sudan adj5 ebola*) or (sudan adj5 filovirus*) or SUDV).ti,ab,kf. | 554 |
| 2 | exp Vaccine/ or Vaccine Development/ or Vaccination/ or vaccin*.ti,ab,kf. | 731,539 |
| 3 | Disease Model/ or exp Enzyme Linked Immunosorbent Assay/ or exp Neutralizing Antibody/ or exp Neutralization Test/ or exp bioassay/ or Enzyme Linked Immunospot Assay/ or (ELISA* or immuno* or immune* or PRNT or neutrali* or monoclonal or mAbs or antiviral* or assay* or luminex or ELISPOT* or T-cell* or (animal adj5 model*) or non-human* or nonhuman* or primate* or chimpanzee* or macaque* or mice or mouse or hamster* or ferret* or murine or guinea pig* or challenge or natural history).ti,ab. or (study or trial).ti. | 10,846,339 |
| 4 | 1 and (2 or 3) | 412 |
| 5 | exp Review/ | 3,418,829 |
| 6 | 4 not 5 | 375 |
| 7 | limit 6 to (english language and "remove medline records") | 84 |

#### Europe PMC

Restricted to preprints

((TITLE:"sudanvirus*") OR (ABSTRACT:"sudanvirus*") OR (TITLE:"SUDV") OR (ABSTRACT:"SUDV") OR ((TITLE:"sudan") AND ((TITLE:"ebola*") OR (TITLE:"virus*")))) AND (SRC:PPR) = 35

#### Ebolavirus (EBOV)

Modifications were made to the MARV/SUDV search because of the considerably larger volume of literature for EBOV. (Without modifications, the search retrieved over 6000 records in PubMed alone.) The impact of the following changes was assessed to minimise the chance of excluding potentially relevant studies.

**Changes to the PubMed EBOV search**

| | |
|---|---|
| 1 | Removed MeSH terms with fewer than 3 hits from the MARV included studies: Biological Assay[mh], Enzyme-Linked Immunospot Assay[mh] OR PRNT[tiab], luminex[tiab], ELISPOT*[tiab] |
| 2 | Replaced Vaccines[mh] with Ebola Vaccines[mh] |
| 3 | Restricted ebola*[tiab] OR EBOV[tiab] to non-MEDLINE indexed records. MEDLINE indexed records limited to ebola[ti] and EBOV[ti] only.<br><br>Restricted vaccin*[tiab] to non-MEDLINE indexed records. MEDLINE indexed records limited to vaccin*[ti] only. |
| 4 | Replaced challenge[tiab] with "animal challenge"[tiab] OR "lethal challenge"[tiab] OR "human challenge"[tiab] OR "infection challenge"[tiab] |
| 5 | Removed study[ti] OR trial[ti] |
| 6 | Excluded Review[pt] OR News[pt] OR Editorial[pt] |
| 7 | Added Filoviridae[mh:noexp] OR Filoviridae Infections[mh:noexp] |

The Embase search was modified in a similar way to the PubMed strategy, plus the removal of conference abstracts, restriction to English language, and searching ebola / EBOV as free-text terms in the title and author keywords only. The Emtree terms Ebolavirus/ or Ebola Hemorrhagic Fever/ were searched as focused terms.

We conducted a separate search for systematic reviews of EBOV (see below). We decided not to search Europe PMC for EBOV because Embase already includes preprints from 2020 onwards from the following sources: bioRxiv, medRxiv, Research Square and SSRN.

Search date: 20 January 2025

**PubMed**

| # | Concept | Search statement | Results |
|---|---|---|---|
| 1 | MeSH Terms for Ebola and filovirus (not exploded) and free-text terms for Ebola limited to non-MEDLINE subset | Filoviridae[mh:noexp] OR Filoviridae Infections[mh:noexp] OR Ebolavirus[mh] OR Hemorrhagic Fever, Ebola[mh] OR ebola*[ti] OR EBOV[ti] OR ((ebola*[tiab] OR EBOV[tiab]) NOT medline[sb]) | 10,314 |
| 2 | MeSH Terms and free-text terms (limited to non-MEDLINE subset) for vaccines | Ebola Vaccines[mh] OR Vaccine Development[mh] OR Vaccination[mh] OR vaccin*[ti] OR (vaccin*[tiab] NOT medline[sb]) | 191,942 |
| 3 | MeSH Terms and free-text terms for immunobridging strategy, in-vitro, human challenge, animal models, natural history | Disease Models, Animal[mh] OR Enzyme-Linked Immunosorbent Assay[mh] OR Antibodies, Neutralizing[mh] OR Antibodies, Monoclonal[mh] OR Neutralization Tests[mh] OR Immunity[mh] OR ELISA*[tiab] OR immuno*[tiab] OR immune*[tiab] OR neutrali*[tiab] OR monoclonal[tiab] OR mAbs[tiab] OR antiviral*[tiab] OR assay*[tiab] OR T-cell*[tiab] OR "animal model"[tiab:~5] OR "animal models"[tiab:~5] OR non-human*[tiab] OR nonhuman*[tiab] OR primate*[tiab] OR chimpanzee*[tiab] OR macaque*[tiab] OR mice[tiab] OR mouse[tiab] OR hamster*[tiab] OR ferret*[tiab] OR murine[tiab] OR "guinea pig*"[tiab] OR "animal challenge"[tiab] OR "lethal challenge"[tiab] OR "human challenge"[tiab] OR "infection challenge"[tiab] OR "natural history"[tiab] | 6,145,497 |
| 4 | Combine sets | #1 AND (#2 OR #3) | 3986 |
| 5 | Exclude reviews, etc. | Review[pt] OR News[pt] OR Editorial[pt] | 4,369,720 |
| 6 | Combine sets | #4 NOT #5 | 3195 |

**Ovid Embase**

Embase Classic + Embase <1947 to 2025 January 17>

| # | Search Statement | Results |
|---|---|---|
| 1 | exp *Ebolavirus/ or *Ebola Hemorrhagic Fever/ or (ebola* or EBOV).ti,kf | 9975 |
| 2 | Ebola Vaccine/ or Vaccine Development/ or Vaccination/ or vaccin*.ti,ab,kf. | 622,276 |
| 3 | Disease Model/ or exp Enzyme Linked Immunosorbent Assay/ or exp Neutralizing Antibody/ or exp Neutralization Test/ or (ELISA* or immuno* or immune* or neutrali* or monoclonal or mAbs or antiviral* or assay* or T-cell* or (animal adj5 model*) or non-human* or nonhuman* or primate* or chimpanzee* or macaque* or mice or mouse or hamster* or ferret* or murine or guinea pig* or (animal or lethal or human or infection adj1 challenge) or natural history).ti,ab. | 7,659,131 |
| 4 | 1 and (2 or 3) | 4373 |
| 5 | exp Review/ or (Editorial or Note or Conference Abstract).pt. | 10,456,88 |
| 6 | 4 not 5 | 3172 |
| 7 | limit 6 to (english language and "remove medline records") | 511 |

Search strategy for systematic reviews of Ebolavirus (limited to PubMed only)

Search date: 13 February 2025

(Filoviridae[mh:noexp] OR Filoviridae Infections[mh:noexp] OR Ebolavirus[mh] OR Hemorrhagic Fever, Ebola[mh] OR ((ebola*[tiab] OR EBOV[tiab]) NOT medline[sb])) AND (Ebola Vaccines[mh] OR Vaccine Development[mh] OR Vaccination[mh] OR (vaccin*[tiab] NOT medline[sb]) OR Disease Models, Animal[mh] OR Enzyme-Linked Immunosorbent Assay[mh] OR Antibodies, Neutralizing[mh] OR Antibodies, Monoclonal[mh] OR Neutralization Tests[mh] OR Immunity[mh] OR ELISA*[tiab] OR immuno*[tiab] OR immune*[tiab] OR neutrali*[tiab] OR monoclonal[tiab] OR mAbs[tiab] OR antiviral*[tiab] OR assay*[tiab] OR T-cell*[tiab] OR "animal model"[tiab:~5] OR "animal models"[tiab:~5] OR non-human*[tiab] OR nonhuman*[tiab] OR primate*[tiab] OR chimpanzee*[tiab] OR macaque*[tiab] OR mice[tiab] OR mouse[tiab] OR hamster*[tiab] OR ferret*[tiab] OR murine[tiab] OR "guinea pig*"[tiab] OR "animal challenge"[tiab] OR "lethal challenge"[tiab] OR "human challenge"[tiab] OR "infection challenge"[tiab] OR "natural history"[tiab]) AND (systematic[sb] OR meta-analysis[pt] OR "systematic review"[tiab] OR meta-analysis[tiab] OR review[ti] OR synthesis[ti])

Retrieved 127 records.

#### Bundibugyo (BDBV)

Date of searches: 18 May 2026

##### PubMed

| # | Bundibugyo virus | Search statement | Results |
|---|---|---|---|
| 1 | Title/abstract terms for BDBV | bundibugyo*[tiab] OR BDBV[tiab] OR BEBOV[tiab] OR BDBOV[tiab] | 202 |
| 2 | MeSH Terms and title/abstract terms for vaccine | (Vaccines[mh] OR Vaccine Development[mh] OR Vaccination[mh] OR vaccin*[tiab] | 566,485 |
| 3 | MeSH Terms and free-text terms for immunobridging strategy, in-vitro, human challenge, animal models, natural history | Disease Models, Animal[mh] OR Enzyme-Linked Immunosorbent Assay[mh] OR Antibodies, Neutralizing[mh] OR Antibodies, Monoclonal[mh] OR Neutralization Tests[mh] OR Biological Assay[mh] OR Enzyme-Linked Immunospot Assay[mh] OR Immunity[mh] OR ELISA*[tiab] OR immuno*[tiab] OR immune*[tiab] OR PRNT[tiab] OR neutrali*[tiab] OR monoclonal[tiab] OR mAbs[tiab] OR antiviral*[tiab] OR assay*[tiab] OR luminex[tiab] OR ELISPOT*[tiab] OR T-cell*[tiab] OR "animal model"[tiab:~5] OR "animal models"[tiab:~5] OR non-human*[tiab] OR nonhuman*[tiab] OR primate*[tiab] OR chimpanzee*[tiab] OR macaque*[tiab] OR mice[tiab] OR mouse[tiab] OR hamster*[tiab] OR ferret*[tiab] OR murine[tiab] OR "guinea pig*"[tiab] OR challenge[tiab] OR "natural history"[tiab] OR study[ti] OR trial[ti] | 9,231,798 |
| 4 | Combine sets | #1 AND (#2 OR #3) | 152 |

##### Embase

Not limited by publication type or language, except removal of ClinicalTrials.gov records.

**Embase Classic + Embase <1947 to 2026 May 14>**

| # | Search Statement | Results |
|---|---|---|

| | | |
|---|---|---|
| 1 | Bundibugyo Ebolavirus/ or (bundibugyo* or BDBV or BEBOV or BDBOV).ti,ab,kf. | 269 |
| 2 | exp Vaccine/ or Vaccine Development/ or Vaccination/ or vaccin*.ti,ab,kf. | 818,080 |
| 3 | Disease Model/ or exp Enzyme Linked Immunosorbent Assay/ or exp Neutralizing Antibody/ or exp Neutralization Test/ or (ELISA* or immuno* or immune* or neutrali* or monoclonal or mAbs or antiviral* or assay* or T-cell* or (animal adj5 model*) or non-human* or nonhuman* or primate* or chimpanzee* or macaque* or mice or mouse or hamster* or ferret* or murine or guinea pig* or (animal or lethal or human or infection adj1 challenge) or natural history).ti,ab. or (study or trial).ti. | 11,606,470 |
| 4 | 1 and (2 or 3) | 207 |
| 5 | Clinical Trial.pt. | 583,802 |
| 6 | 4 not 5 | 204 |
| 7 | limit 6 to "remove medline records" | 56 |

#### Screening

| | Total retrieved | Duplicates removed (EndNote/Covidence) | Total to screen |
|---|---|---|---|
| PubMed | 152 | 0 | 152 |
| Embase | 56 | 10 | 46 |
| Totals | 208 | 10 | 198 |

#### PRISMA flow diagrams for each pathogen evidence map

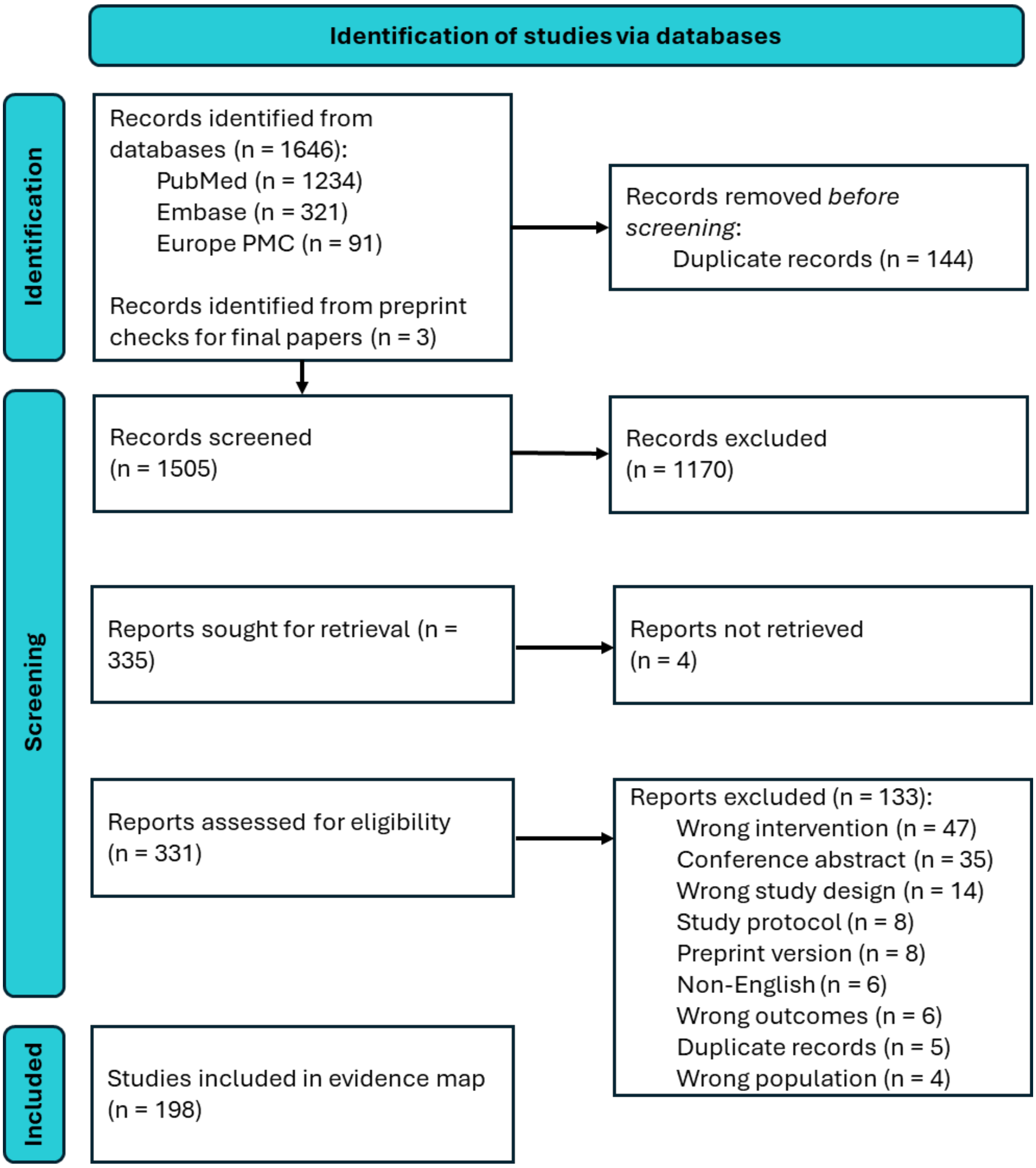

**PRISMA flow diagram for MARV evidence map**

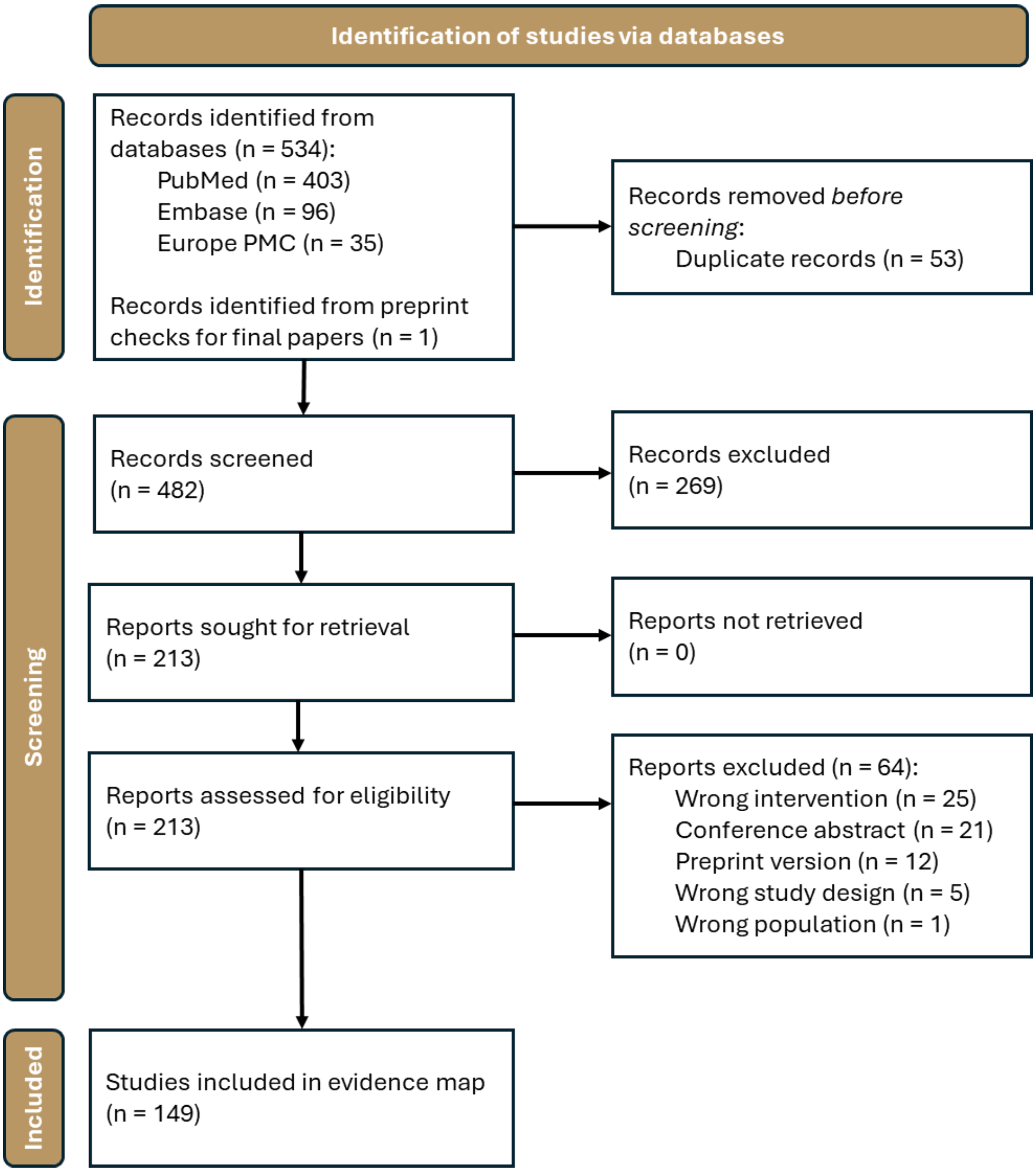

***PRISMA flow diagram for SUDV evidence map***

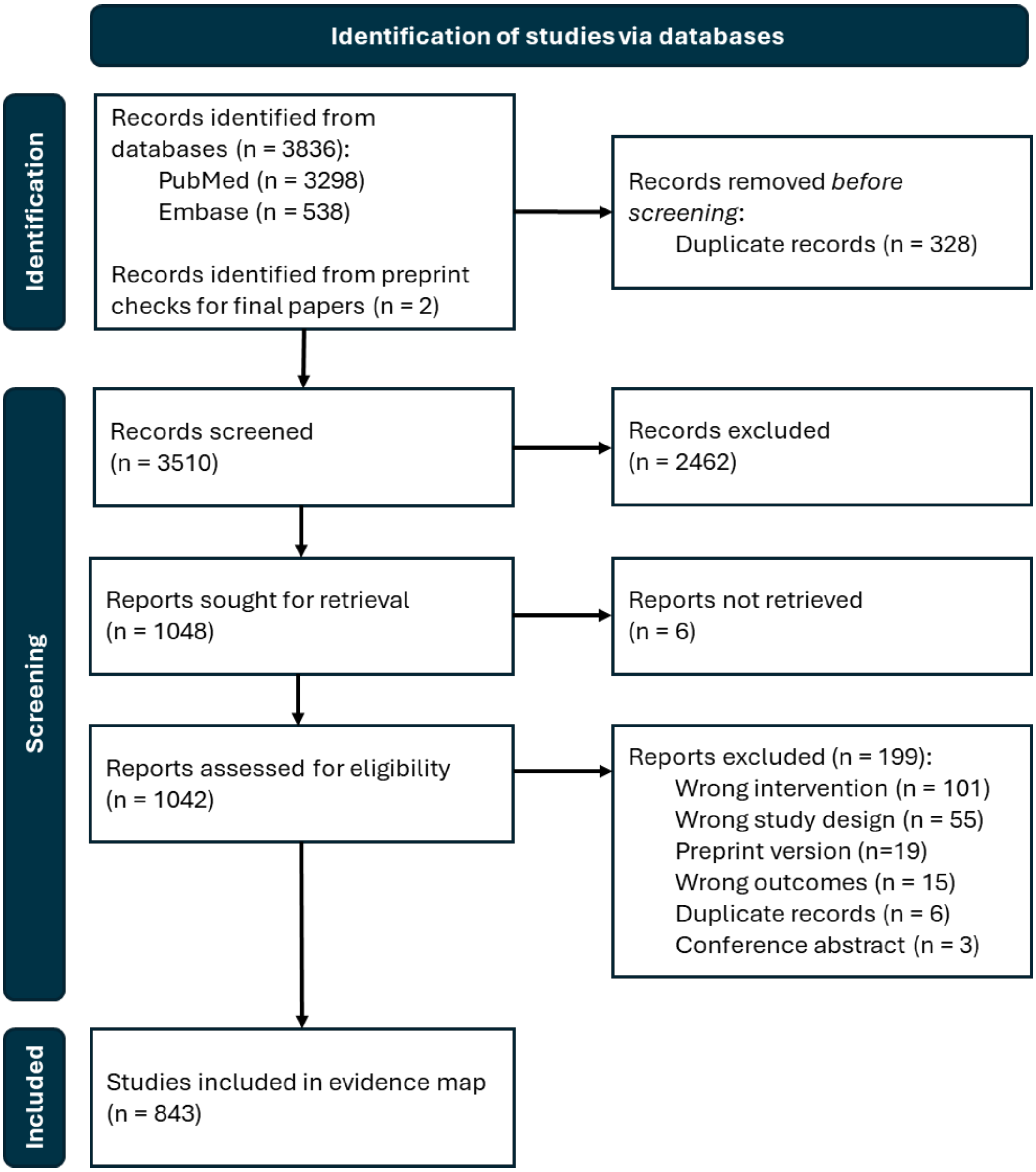

***PRISMA flow diagram for EBOV evidence map***

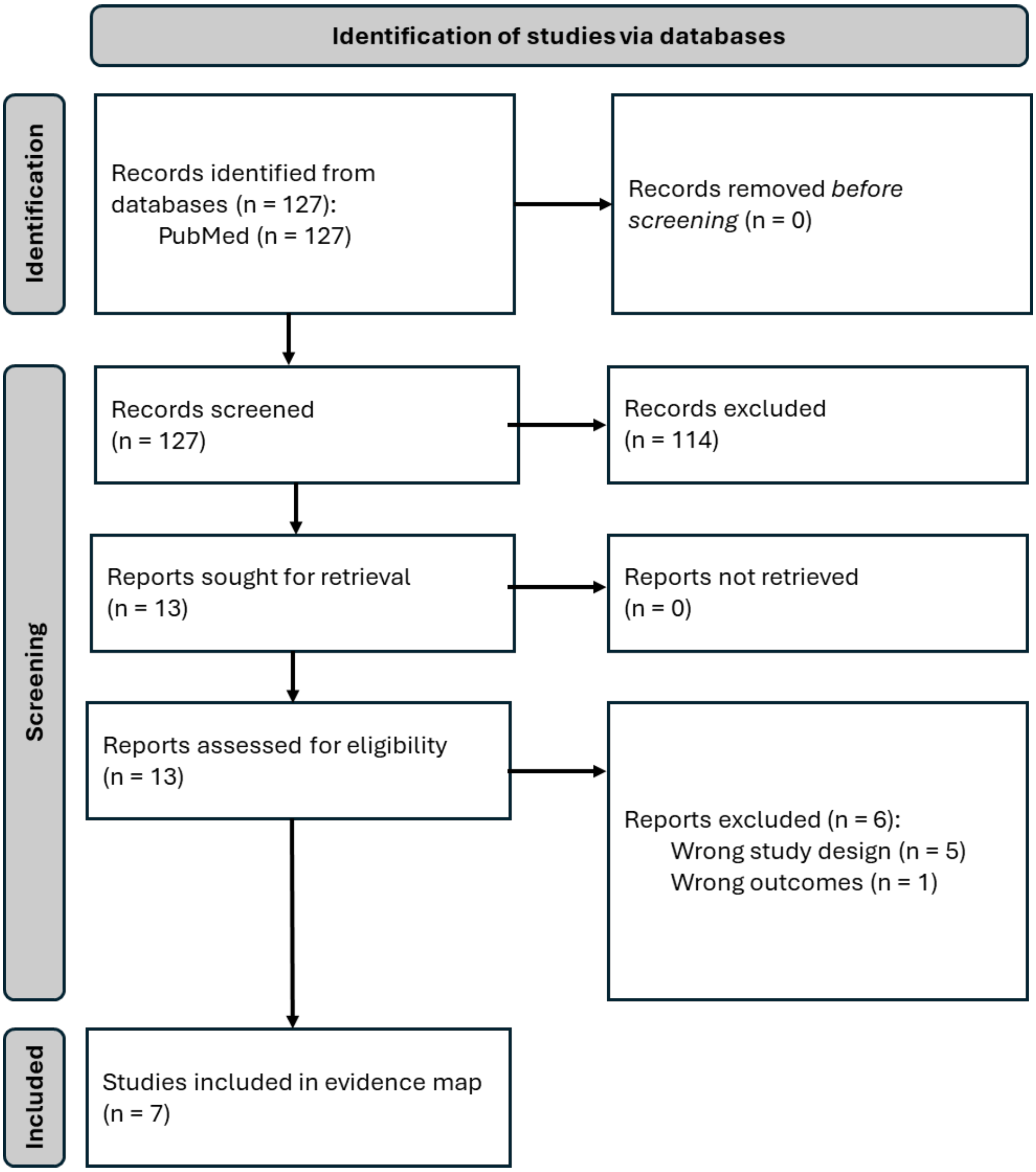

***PRISMA flow diagram for systematic reviews for EBOV evidence map***

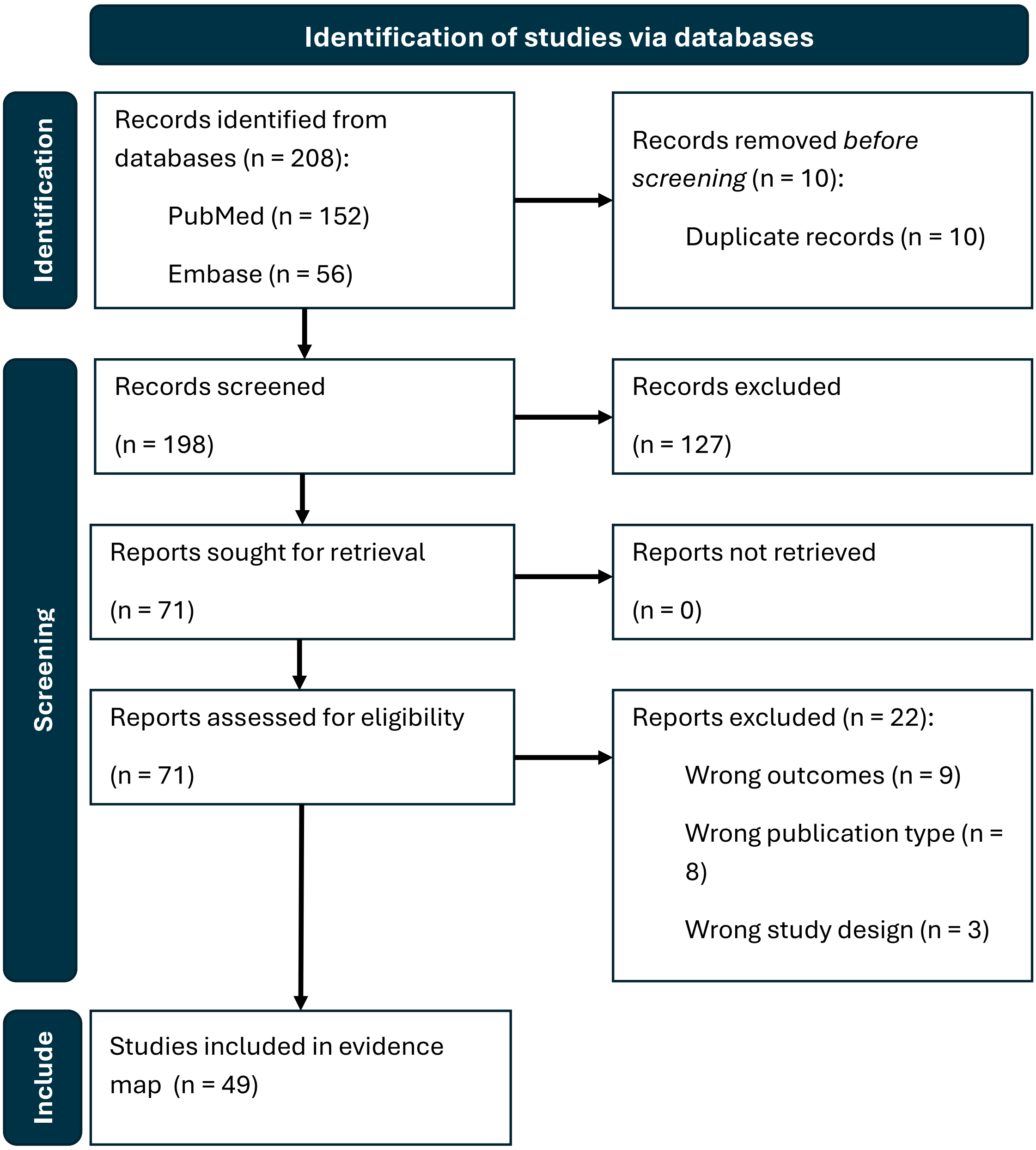

***PRISMA flow diagram for systematic reviews for BDBV***

#### Study Tagging Decision tree for data extractors

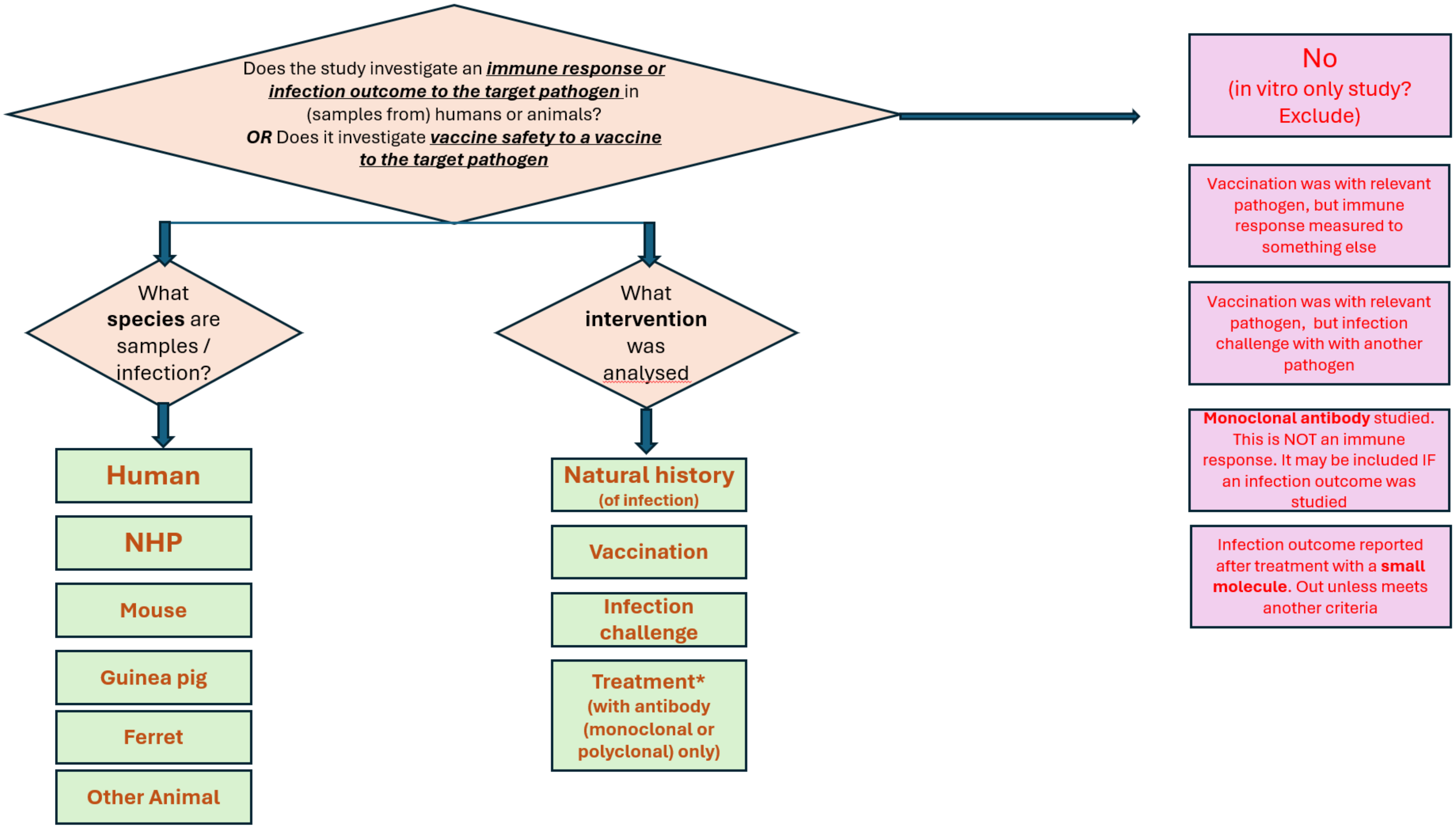

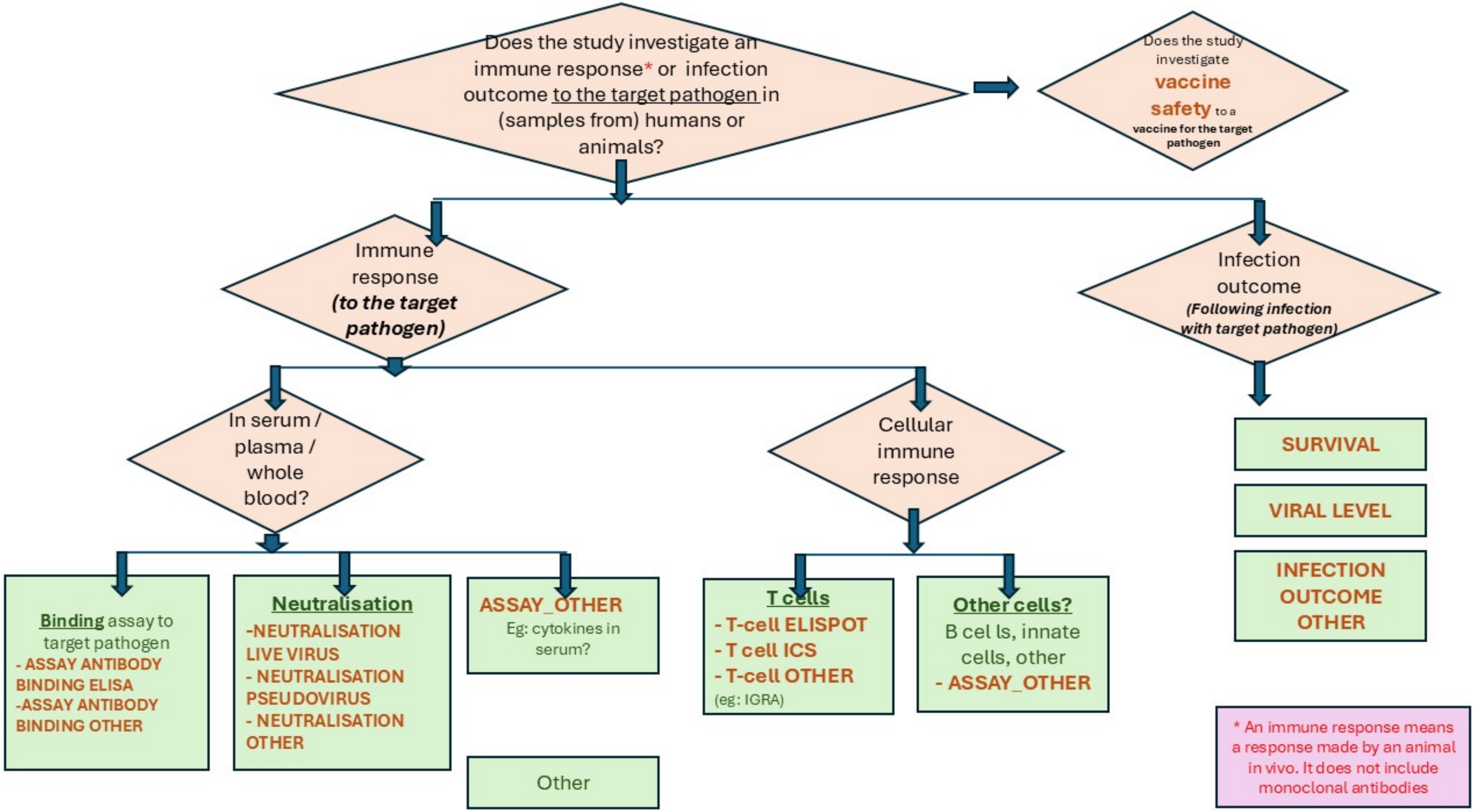
Does the study investigate an immune response* or infection outcome to the target pathogen in (samples from) humans or animals?
Does the study investigate vaccine safety to a vaccine for the target pathogen
Immune response (to the target pathogen)
Infection outcome (Following infection with target pathogen)
In serum / plasma / whole blood?
Cellular immune response
SURVIVAL
VIRAL LEVEL
INFECTION OUTCOME OTHER
Binding assay to target pathogen
- ASSAY ANTIBODY BINDING ELISA
-ASSAY ANTIBODY BINDING OTHER
Neutralisation
-NEUTRALISATION LIVE VIRUS
- NEUTRALISATION PSEUDOVIRUS
- NEUTRALISATION OTHER
ASSAY_OTHER
Eg: cytokines in serum?
Other
T cells
- T-cell ELISPOT
- T cell ICS
- T-cell OTHER
(eg: IGRA)
Other cells?
B cel ls, innate cells, other
- ASSAY_OTHER
* An immune response means a response made by an animal in vivo. It does not include monoclonal antibodies
